# Real-time molecular classification of leukemias

**DOI:** 10.1101/2022.06.22.22276550

**Authors:** Mélanie Sagniez, Shawn M. Simpson, Maxime Caron, Marieke Rozendaal, Bastien Paré, Thomas Sontag, Sylvie Langlois, Alexandre Rouette, Vincent-Philippe Lavallée, Sonia Cellot, Daniel Sinnett, Thai Hoa Tran, Martin A. Smith

**Author notes:** Contributed equally (Mélanie Sagniez, Shawn M. Simpson).

## Abstract

Gene expression profiling provides a detailed molecular snapshot of cellular phenotypes that can be used to compare different biological conditions. Nanopore sequencing technology can generate high-resolution transcriptomic data in real-time and at low cost, which heralds new opportunities for molecular medicine. In this study, we demonstrate the clinical utility of real-time transcriptomic profiling by processing RNA sequencing data from childhood acute lymphoblastic leukemia (ALL) patients on-the-fly with a trained neural network classifier. This strategy successfully distinguished 11/12 representative ALL molecular subtypes and one non-leukemia control in as little as 5 minutes of sequencing on a *MinION* sequencer or in less than 1 hour on disposable, low cost *Flongle* flow cells. Our findings suggest that real-time transcriptomics constitutes a drastically efficient solution for the molecular diagnosis of ALL and other diseases, where conventional clinical workflows require days if not weeks to achieve similar results.

## Introduction

Acute leukemia is the most common pediatric cancer accounting for over 1/3 of cases in children and young adults, of which acute lymphoblastic leukemia (ALL) will affect 4 out of 5 leukemia patients *(1)*. Survival outcomes in childhood ALL have significantly improved over the past 5 decades mainly due to refined, risk-adapted treatment intensification based on patients’ presenting features, leukemia cytogenetics, minimal residual disease and an improved understanding of ALL biology. The molecular landscape of ALL is characterized by diverse genetic alterations that define novel molecular subtypes, which confer prognosis and unravel tailored therapeutic vulnerabilities *(2)*. Nevertheless, the outcome of adolescents and young adults with ALL between 16-30 years of age remains significantly inferior compared to their younger counterparts due to higher rates of both relapses and treatment-related mortality. Moreover, ALL is an aggressive disease and can present with life-threatening symptoms at diagnosis, thus substantiating the need for rapid molecular diagnostics.

The current gold standard for ALL diagnosis, classification and risk stratification involves bone marrow morphology and flow cytometry, followed by conventional cytogenetics and molecular assays to identify recurrent genetic alterations. This process encompasses several tests, including G-band karyotyping, fluorescence *in situ* hybridization (FISH), comparative genome hybridization (CGH) and reverse-transcription polymerase chain reaction (RT-PCR) which are labor-cumbersome, time-consuming and require multiple samples.

The advent of next-generation sequencing (NGS), specifically RNA-sequencing (RNA-seq), has expanded ALL taxonomy from 7 conventional to 19 molecular ALL subtypes, which are defined by gene alterations and/or distinct gene expression signatures; features that are either cryptic or not detected by conventional diagnostic pipelines *(3)*. Therefore, several groups are now incorporating RNA-seq into clinical workflows to refine the molecular classification and risk stratification of ALL *(3)*. However, NGS is currently hampered by a relatively long turnaround time for decision-making (typically 3 to 4 weeks), remains a costly endeavor and is only available in limited institutions.

Third-generation (real-time, single molecule) sequencing technologies offer unique possibilities in this space. Portable and accessible nanopore sequencers, such as the Oxford Nanopore Technologies’ (ONT) MinION device, yield impressive throughput and low error-rates, the latter situated below 5%. Given the speed and accessibility of nanopore sequencing, we hypothesized that processing RNA-seq data from leukemia patients in real-time would improve the turnaround time for disease subtype classification. Here, we leverage a neural network classifier trained on heterogenous short-read sequencing data to demonstrate how nanopore RNA-seq can generate diagnostic-grade molecular classification of ALL patients as fast as 5 minutes of sequencing for a fraction of the cost of contemporary clinical workflows.

## Materials and methods

### Sample cohort

Study samples consist of 12 ALL patients (11 pre-B and 1 pre-T) from the established Quebec childhood ALL (QcALL) cohort *(4)* and one control sample. Patients (8 boys and 4 girls), aged from 1 to 17 years (median 10 years) were diagnosed in the Division of Pediatric Hematology-Oncology at Sainte-Justine University Health Center (UHC), Montreal, Canada, between 2017 and 2021. The Sainte-Justine Institutional Review Board approved the research protocol, and written informed consent was obtained from all participating individuals or their parents (see Human Research Considerations section).

### Training set and classifier

The training set used is described by Tran et al. *(3)* and contains 1,134 ALL RNA-seq samples generated with diverse short-read sequencing protocols and platforms. These include both in-house (n=72) and public datasets (n=1,062) *(5, 6)* that are distributed amongst 17 well-documented ALL subtypes. The data were corrected for batch effects using a Surrogate Variable Analysis (SVA) package *(7)* and the 500 most differentially expressed genes obtained from the DESeq2 likelihood ratio test (LRT) *(8)*, which were used to train a single layer neural network classifier in Tran et al. *(3)*. As some samples in the test set might also be present in the training set for the in-house samples, we ensured that the latter were removed from the training set when spawning the neural network to avoid overfitting.

### Sample preparation and nanopore sequencing

Library preparation was done on RNA isolated from either frozen or fresh patient samples using TRIzol and following the manufacturer’s protocol (Invitrogen, Mississauga, ON, Canada). Different protocols were used based on the different RNA-seq chemistries and devices tested. The tested devices were MinION cDNA, MinION dRNA and MinION Flongle. RNA concentrations were obtained using a Qubit machine and the Qubit RNA assay kit (high sensitivity) [Invitrogen, Mississauga, ON, Canada], while post-PCR cDNA was quantified using the Qubit 1X dsDNA BR Assay Kit (Invitrogen, Mississauga, ON, Canada). When required, the sequencing runs were refueled with 250 uL of FB buffer (with FLT) [Oxford Nanopore Technologies, Oxford, UK].

#### MinION cDNA preparation protocol

All cDNA samples sequenced on a MinION flowcell (R9.4.1 - FLO-MIN106) [Oxford Nanopore Technologies, Oxford, UK] were prepared following manufacturer’s protocol (SQK-PCS109) [Oxford Nanopore Technologies, Oxford, UK]. PCR was done as follows: 95°C for 30 seconds for 1 cycle (initial denaturation), 95°C for 15 seconds for 15 cycles (denaturation), 62°C for 15 seconds for 15 cycles (annealing), 65°C for 5 minutes for 15 cycles (extension), 65°C for 10 minutes for 1 cycle (final extension). Final libraries were primed on the flowcells following manufacturer’s recommendations and ran for 72 hours on a GridION sequencer (samples 0205, 0215, 0232, 0236, 0243, 0503) or 89 hours (sample 0318)

#### MinION dRNA preparation protocol

All RNA samples sequenced on a MinION flowcell (R9.4.1 - FLO-MIN106) were prepared following manufacturer’s protocol for dRNA sequencing (SQK-RNA002) [Oxford Nanopore Technologies, Oxford, UK]. Final libraries were loaded on flowcells following manufacturer’s recommendations and ran for 46 hours on a GridION sequencer (samples 0205, 0215, 0232, 0236 and 0243) or 72 hours (sample 0318).

#### Flongle cDNA preparation protocol

All cDNA samples sequenced on a Flongle flow cells (R9.4.1 - FLO-FLG001) [Oxford Nanopore Technologies, Oxford, UK] were prepared following manufacturer’s protocol (SQK-PCS109) [Oxford Nanopore Technologies, Oxford, UK]. PCR was done as follows: 95°C for 30 seconds for 1 cycle (initial denaturation), 95°C for 15 seconds for 15 cycles (denaturation), 62°C for 15 seconds for 15 cycles (annealing), 65°C for 5 minutes for 15 cycles (extension), 65°C for 10 minutes for 1 cycle (final extension). PCR reactions were then processed using SQK-LSK110 library preparation kit, following manufacturer’s recommendations (Oxford Nanopore Technologies, Oxford, UK). Final libraries were primed on the flowcells following manufacturer’s recommendations and ran for 36 hours on a GridION sequencer (samples 0236, 0243, 1018 and 0244), for 20 to 25 hours (samples 0205, 0232, 0503, 0258 and 0274), for 10 hours (sample 0998) or 6 hours (sample 0340). Running time was based on the number of pores available on the flow cell (**Supplementary Table 1**).

### Sequencing data processing

Raw data were base called with Guppy (see **Supplementary Table 1** for software versions) and retrospectively grouped into timepoints based on their acquisition time during sequencing, as reported in raw .fast5 signal files and the base calling output file “sequencing_summary.txt”. For each timepoint, reads were aligned to the Ensembl 75 transcriptome reference (http://feb2014.archive.ensembl.org/Homo_sapiens/Info/Index) and the resulting count matrices were passed to the classification algorithm to output classification probabilities for each time point.

## Results

RNA extracted from bone marrow aspirates of 13 pediatric ALL patients with well-defined molecular subtype classification for ONT sequencing using various chemistries and flow cells (**Supplementary Table 1)**. Samples were selected from the QcALL cohort to represent the diversity of ALL molecular subtypes (**Table 1**). Following completion of the sequencing runs, we grouped the sequencing reads into bins corresponding to various acquisition time intervals; from 60 seconds of sequencing to 4 hours–in addition to the complete (or ‘final’) dataset–to retrospectively emulate real-time data acquisition. Each time point was processed by aligning reads to a reference transcriptome (see Methods) to generate a count matrix, which was then submitted to a neural network classifier pre-trained on ALL RNA-seq data *(3)*.

**Table 1:**
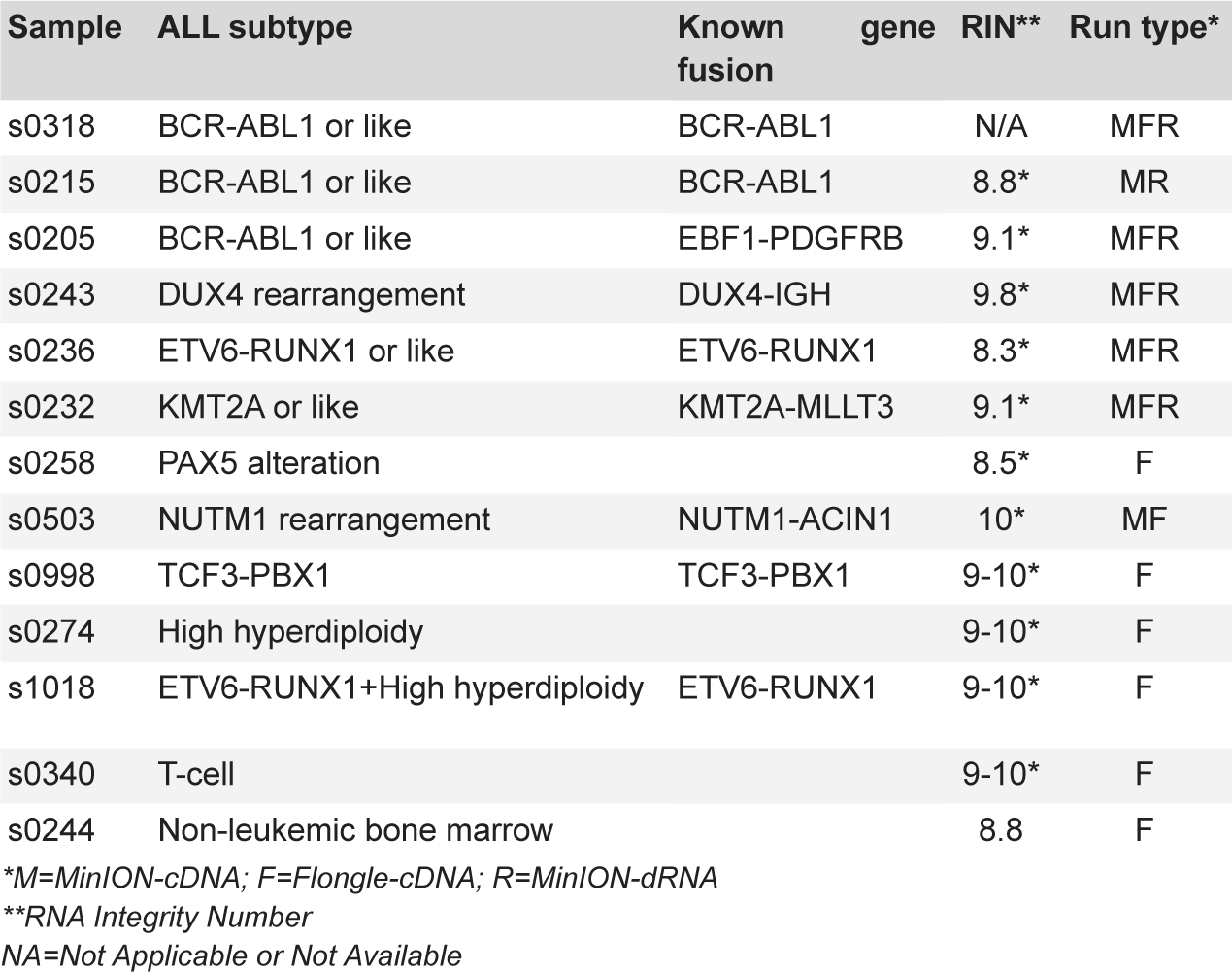
Patient samples and ALL subtypes.

We first performed cDNA sequencing of 7 ALL samples on MinION flow cells, which typically produce 5-15 million reads over 48 hours. All but one sample was assigned to the correct molecular ALL subtype in less than 5 minutes of sequencing with >90% probability (**Figure 1**). Sample s0236–a B-ALL patient with ETV6-RUNX1 fusion, the most common pediatric ALL gene fusion, produced the fastest classification; 3 minutes of sequencing were sufficient to obtain a 98% molecular classification probability. A classification of 99% was obtained for subsequent time points. Two BCR-ABL1 or BCR-ABL1-like samples, s0205 and s0318, as well as s0232 (a KMT2A fusion) required only 4 minutes of sequencing to achieve 96%, 97% and 94% classification probability, respectively, while sample s0215 required 5 minutes to achieve 95%. Of the remaining samples, s0243 (a DUX4 fusion) attained 92% classification probability in 5 minutes but dipped to ∼80% on 2 occasions before stabilizing towards 98% probability after 2 hours of sequencing. Based on these 6 samples, 5 minutes is enough to accurately classify ALL subtypes using MinION flow cells and cDNA on Nanopore sequencers. The classifier failed to correctly classify s0503, a rare NUTM1 fusion. However, the NUTM1-rearranged molecular subtype was predicted at the end of the sequencing run (after 72 hours of sequencing) with a probability of 37%, albeit BCR-ABL1 and PAX5 alteration classifications also produced 17% and 34% classification probabilities, respectively.

**Figure 1:**
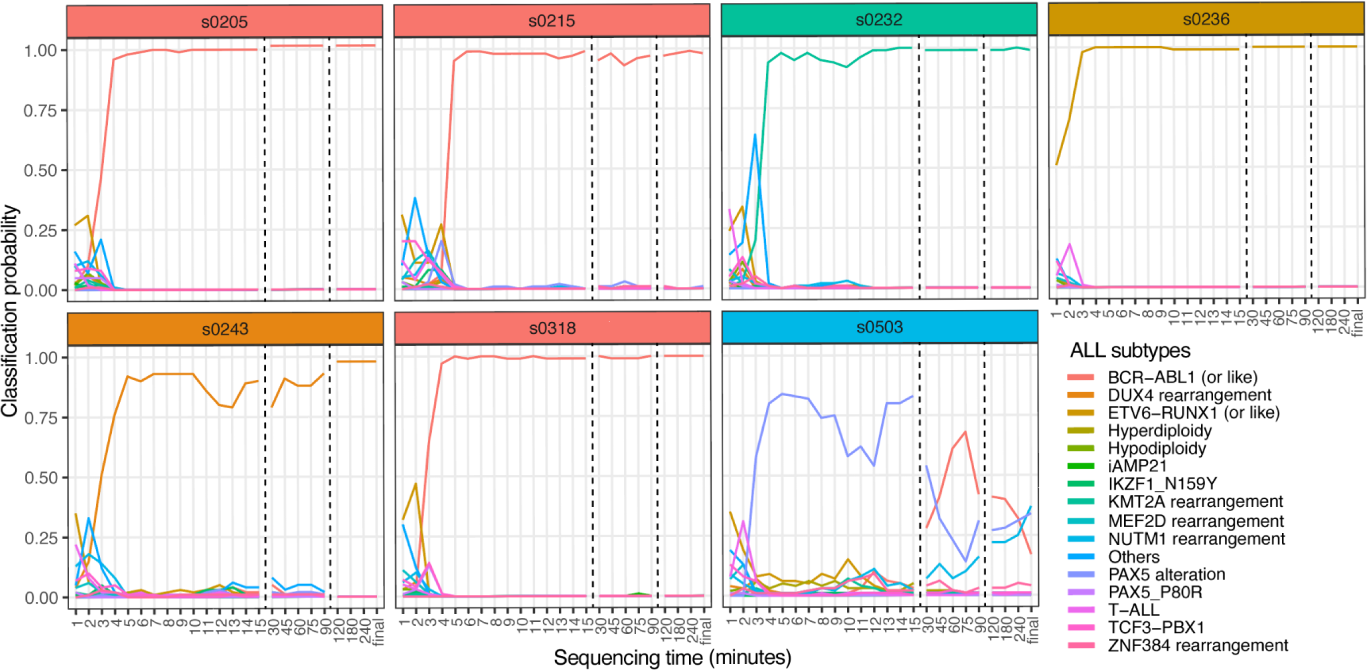
Molecular subtype classification from cDNA reads on a MinION flow cell in function of sequencing time. Independent clinical classification is indicated by header colors for each sample.

Given the speed of this approach with MinION flow cells, we wondered if similar classification results could be achieved on lightweight and disposable Flongle flow cells, which generate ∼15x less data than MinION flow cells. As expected, more time was required to obtain high confidence ALL subtype classification probabilities on Flongle chips (**Figure 2**), which nonetheless yielded accurate classifications with probabilities >90% in less than 1 hour for 9 out of 11 samples. Interestingly, sample s0243 was processed before the formal clinical classification was received, initially being assigned the subtype “other”. However, our classifier suggested a DUX4 rearrangement subtype with a probability of 95% after 60 minutes of sequencing, which was subsequently confirmed by identification of the gene fusion via short read RNA-seq.

**Figure 2:**
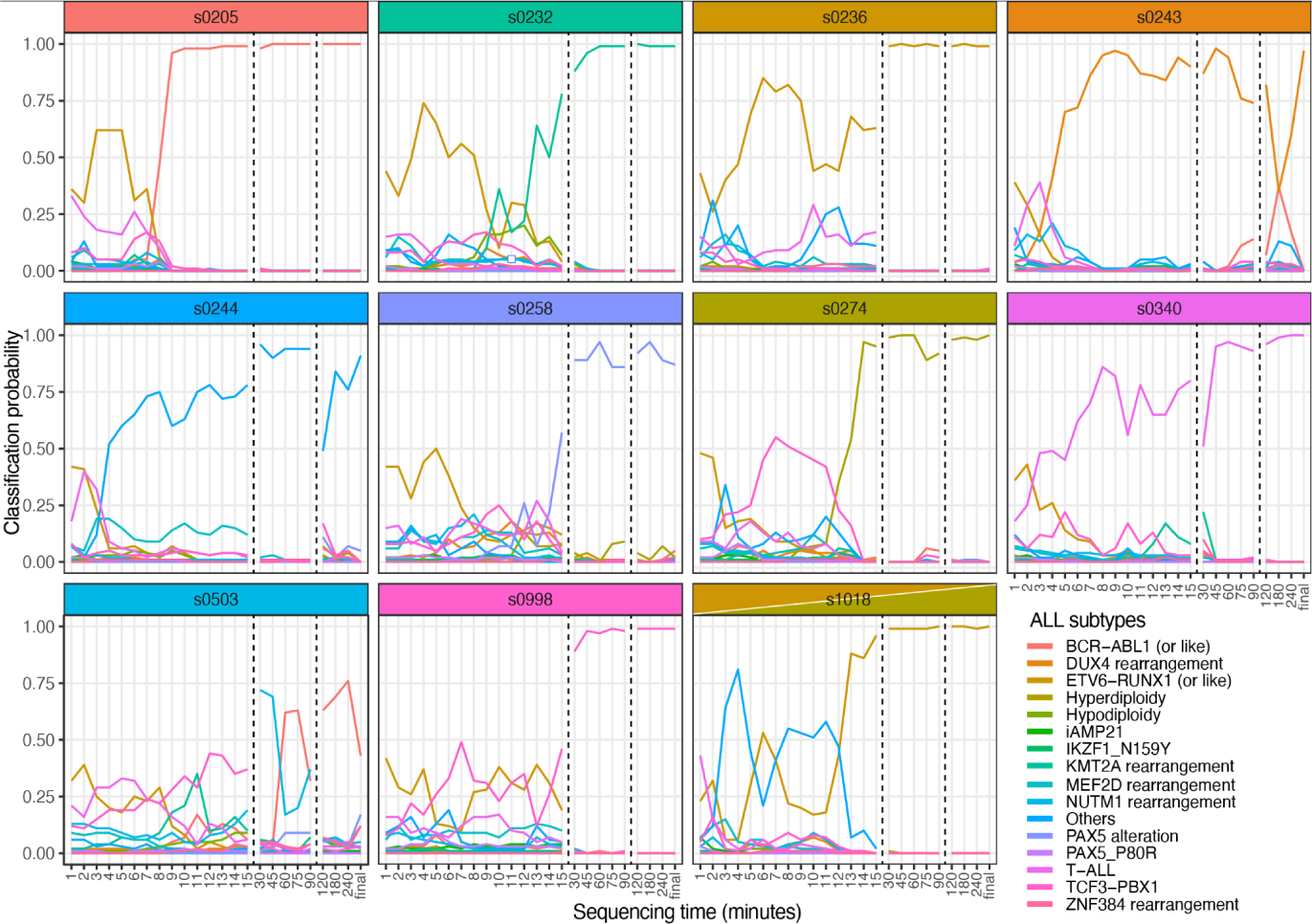
Molecular subtype classification from cDNA reads on Flongle flow cells in function of sequencing time. Sample s1018 is associated with a double clinical classification, as indicated by the header colors.

Two samples subjected to Flongle cDNA sequencing were problematic. The first, s0503 (the above-mentioned NUTM1 rearrangement) was ultimately classified as the BCR-ABL1 or like subtype, consistent with previous results. The second, s1018 (a rare leukemia sample characterized by 2 distinct genomic aberrations; an ETV6-RUNX1 fusion and hyperdiploidy) was accurately classified as ETV6-RUNX1 after 15 minutes of sequencing with a probability of 96%. Nonetheless, the classifier failed to pick up the second subtype associated with this sample that is high hyperdiploidy (0% for all timepoints), which may suggest that the gene fusion is linked to the dominant molecular phenotype in this cancer.

We also included a bone marrow sample from a patient that was not diagnosed with leukemia (s0244) as a negative control. The deterministic nature of the classifier accurately recognized this sample as an “others” subtype, a category that was trained from ALL samples without canonical gene fusions and/or gene expression profiles. Interestingly, the above-mentioned sample with a dual clinical classification was classified as “others” early in the data acquisition.

To assess whether an independent sequencing chemistry could generate comparable results, poly(A) RNA sequencing (no conversion to cDNA and use of PCR) was applied to samples with sufficient quantities of RNA (**Figure 3**). Native RNA sequencing on MinION flow cells produced similar yields and results to Flongle cDNA experiments (**Figure 4**, top panel). The fastest correct classification was for sample s0318 with a 94% probability after 7 minutes of sequencing, a result akin to cDNA-MinION runs. All 6 samples were accurately classified in less than 120 minutes of sequencing with a probability > 90%. A direct comparison of the three different sequencing strategies is presented in **Figure 4**.

**Figure 3:**
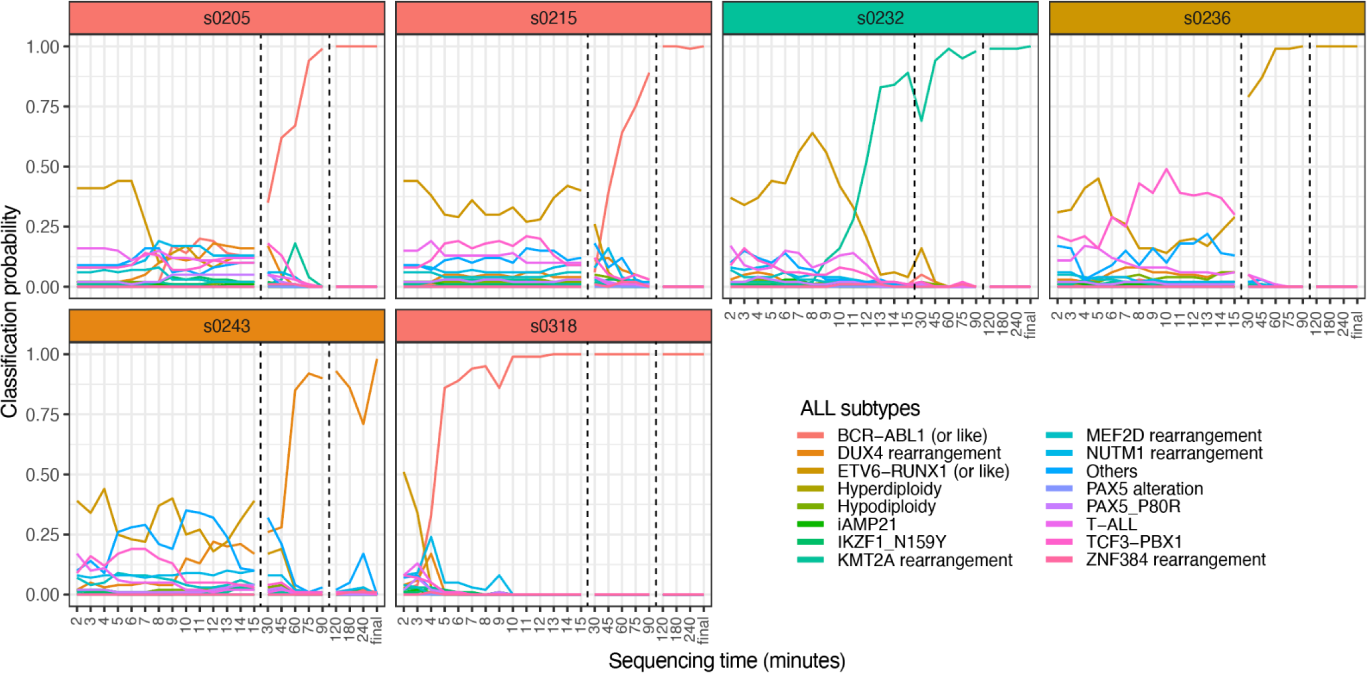
Molecular subtype classification from native RNA reads on MinION flow cells in function of sequencing time.

**Figure 4:**
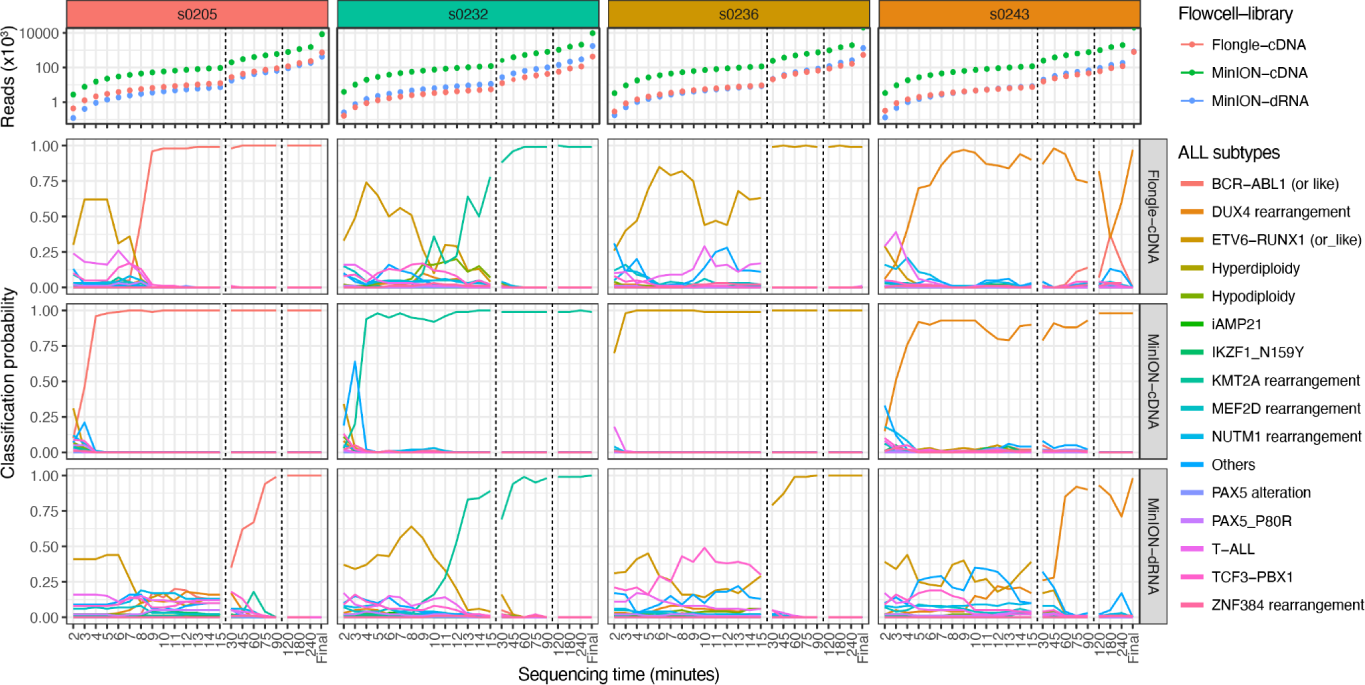
Comparison of time-sensitive molecular subtype classification performance across different nanopore RNA sequencing strategies.

## Discussion

Molecular profiling of acute leukemias via RNA-seq is a powerful tool for the characterization of disease heterogeneity, biomarker discovery and risk stratification of leukemia patients *(2, 9–14*). Our results demonstrate that nanopore sequencing and supervised machine learning can be used to diagnose and accurately classify molecular ALL subtypes in as little as 4 minutes of sequencing, or in ∼4 hours when factoring in RNA extraction and sample preparation time. This constitutes a turnaround time several orders of magnitude faster than current clinical workflows. For most samples, less than 10,000 reads were sufficient to pinpoint the ALL subtype. The slightly longer (1 to 2 hours) sequencing turnaround time required when using Flongle flow cells is nonetheless efficient when considering their significantly lower cost (US$80 versus US$500). This is a significant improvement when considering that contemporary workflows for the molecular classification of ALL samples using conventional cytogenetic/molecular approaches require 2-14 days, cost ∼CA$3000 (without NGS-based RNA-seq), involve multiple different assays and approximately 40% cases remain unclassified.

Overall, we found that cDNA sequencing gave the best results in terms of speed and stability of the predictions over time. The slightly lower efficacy of native RNA sequencing could be explained by the supervised nature of the classifier, which was exclusively trained on previously published Illumina short-read sequencing data from cDNA *(3)*. It would therefore include features derived from reverse transcription and PCR amplification, which would not be present in native RNA data. A similar rationale explains why one sample (s0503, NUTM1 rearrangement) performed poorly on both MinION and Flongle runs; NUTM1 rearrangements represent only 0.5% of the training set (only 6 out of 1,134 samples), including the queried sample that was left out during training. The weak representation of a subtype in the training set might corrupt the neural network and orient the prediction toward well-represented subtypes, such as BCR-ABL1 that are more overrepresented in the training set. Additional patient samples of rarer ALL subtypes, alternative machine learning models and nanopore-specific training data would undoubtedly improve this strategy in the future. A conservative threshold (e.g. 90%) and longer sequencing times would be sufficient to distinguish low-confidence predictions.

The unique sample with dual clinical classification (s1018, ETV6-RUNX1 + high hyperdiploidy) emitted a strong, unique classification probability to the ETV6-RUNX1 subtype only after 30 minutes of sequencing. As another high hyperdiploidy ALL sample (s0274) was correctly classified with strong probability, it is possible that the neural network identified the ETV6-RUNX1-derived gene expression signature as the predominant molecular phenotype of this sample. Additional molecular studies would be required to validate this hypothesis. Interestingly, the classification from early timepoints (i.e shallow sequencing data) assigned this sample to the “others” ALL subtype, which was also the category assigned to the only negative control used in this study (a bone marrow sample from a patient without leukemia). Therefore, this model cannot distinguish between atypical leukemias and non-leukemias in its current implementation. More leukemia and control samples are thus required to define new subtype categories and to validate the clinical utility and precision of our approach.

Two pediatric ALL classification tools based on gene expression signatures were published while preparing this manuscript. The first, ALLSorts is a B-ALL classifier that performs hierarchical supervised classification within 5 meta-subtypes (Ph, KMT2A, ETV6-RUNX1, ZNF384 and High Ploidy Signature groups) and 18 subtypes pretrained on 1,223 B-ALL samples *(15)*. As for the second, after comparing multiple machine-learning classification models, the selected strategy employs a partial least-squares discriminant classifier trained on 1,036 samples from ALL lineages (B and T) and acute myeloid leukemia (AML) genomic subtypes. It produces binary predictions to be used as features that are input into a non-linear Support Vector Machine (SVM) classifier for 3 leukemia lineages and 8 subtypes *(14)*. Our classifier employs a feedforward neural network (FNN) trained on 1,134 ALL samples and outputs prediction probabilities for 16 ALL subtypes. As the FFN wasn’t considered in the Wang et al. study, a comparison between these tools could enhance our results and further reduce the sequencing time required for each chemistry and device tested. Also, current classifiers handle between 8 to 18 subtypes, while up to 23 subtypes have been identified *(5, 16)*. This suggests that further refinement and subtype definition is needed in order to find a consensus for homogeneous classifications.

The molecular classification strategy we present herein could nonetheless be improved. Firstly, the machine learning strategy we applied was trained on gene-level NGS-derived quantification data. Full-length isoforms would provide a more diverse and, presumably, more sensitive training set to refine the classifier. Moreover, using an *ad hoc* reference transcriptome assembled from single molecule sequencing of leukemia samples would provide a bespoke qualitative reference, upon which individual sample would be quantified to remove impertinent reference transcripts and include novel isoforms and long non-coding RNAs that are specific to this condition. Although this would require deeper transcriptomic data and the associated clinical metadata generated from patient samples, it has the potential to identify new biomarkers of disease and, consequently, improve disease classification and risk stratification efficiency. Similarly, generating a molecular subtype classifier based on native RNA sequencing data, which would include RNA modifications, would add an additional level of accuracy to disease subtype modeling. Indeed, several RNA modifications, which are often lost during reverse transcription, are dysregulated in leukemias *(17)*.

Secondly, our method does not directly identify gene fusions that define molecular subtypes and identify targeted therapies *(18, 19)*. Identifying fusion genes remains a challenge due to their (often) low expression levels. In this study, few known fusions were detected with the relatively shallow nanopore sequencing we performed (data not shown). Deeper molecular sampling with larger PromethION flowcells, for example, or targeted strategies (e.g. PCR panels, adaptive sampling, CRISPR enrichment, etc) would be required to identify gene fusions more reliably using nanopore sequencers. Lastly, whilst the processing reported herein occurred retrospectively (for direct temporal comparisons between samples), our pipeline can be implemented in real-time with relative ease, which we have successfully performed in-house. N.B. Default sequencing parameters should be adjusted consequently to produce sequencing files with less than 4000 reads (default parameters) in order to perform ultra-rapid (∼5m) classification, especially with Flongle or native RNA sequencing.

In conclusion, the diagnostic strategy we propose herein successfully leverages real-time nanopore sequencing to provide a tangible solution for ALL diagnostics and risk-stratification, potentially providing same-day recommendations for precision therapeutic strategies. In addition to its rapid turnaround time, the accuracy, cost and portability of nanopore RNA sequencing provides exciting new opportunities for molecular medicine in the post-genomics era.

## Data Availability

All data produced in the present study are available upon reasonable request to the authors.

## Author contributions

MAS conceived the study. MS, SMS and MAS wrote the manuscript with input from all authors. MS, SMS, MC & MAS performed bioinformatics analyses. MS, SMS, MC, performed data analysis. MS, MR, BP, TS, SL performed sample preparation. MS, MR & BP performed nanopore sequencing. AR, VL, SC, DS, THT & MAS supervised, revised manuscript and contributed to experimental design.

## Acknowledgments

This work was partially funded by a Cole Foundation transition award, a Canadian Foundation for Innovation John R. Evans Leaders Fund (40767) and a National Science and Engineering Research Council of Canada Discovery Grant (RGPIN-2022-04265) to MAS and by a *Fondation Charles-Bruneau* grant to DS. MS is partially supported by a Cole Foundation fellowship. BP is partially supported by a *Fonds de Recherche du Québec - Santé* postdoctoral fellowship. MAS is partially supported by a *Fonds de Recherche du Québec - Santé* Junior 1 fellowship and establishment award (307539). We would also like to thank participating patients and their families, as well as the Fondation CHU Sainte-Justine for their global support.

## Human research considerations

This study (project code MP-21-2019-2032) is approved for human research by the CHU Sainte-Justine ethics committee (FWA00021692) designated by the *Ministère de la Santé et des Services Sociaux du Québec*.

## Availability of data and materials

All data produced in the present study are available upon reasonable request to the authors.

